# Hydroxychloroquine in the Pregnancies of Women with Lupus: A Meta-Analysis of Individual Participant Data

**DOI:** 10.1101/2021.12.21.21267285

**Authors:** Megan E. B. Clowse, Amanda M. Eudy, Stephen Balevic, Gillian Sanders-Schmidler, Andrzej Kosinski, Rebecca Fischer-Betz, Dafna Gladman, Yair Molad, Cecilia Nalli, Abir Mokbel, Angela Tincani, Murray Urowitz, Caroline Bay, Megan van Noord, Michelle Petri

**Affiliations:** Duke University; Heinrich-Heine-Universität Düsseldorf; University of Toronto; Rabin Medical Center; Sackler Faculty of Medicine, Tel Aviv University; Spedali Civili and University of Brescia; Cairo University; Baylor College of Medicine; University of California, Davis; Johns Hopkins School of Medicine

**Keywords:** Systemic lupus erythematosus, Pregnancy, Hydroxychloroquine, Pregnancy loss, Preterm birth

## Abstract

**Objective:** Multiple guidelines recommend continuing hydroxychloroquine (HCQ) for systemic lupus erythematosus (lupus) during pregnancy based on observational data. The goal of this individual patient data meta-analysis was to combine multiple datasets to compare pregnancy outcomes in women with lupus on and off HCQ.

**Methods:** Eligible studies included prospectively-collected pregnancies in women with lupus. After a manuscript search, 7 datasets were obtained. Pregnancy outcomes and lupus activity were compared for pregnancies with a visit in the first trimester in women who did or did not take HCQ throughout pregnancy. Birth defects were not systematically collected. This analysis was conducted in each dataset and results were aggregated to provide a pooled odds ratio.

**Results:** Seven cohorts provided 938 pregnancies in 804 women. After selecting one pregnancy per patient with a 1^st^ trimester visit, 668 pregnancies were included; 63% took HCQ throughout pregnancy. Compared to pregnancies without HCQ, those with HCQ had lower rates of highly active lupus, but did not have different rates of fetal loss, preterm birth, or preeclampsia. Among women with low lupus activity, HCQ reduced the risk for preterm delivery.

**Conclusion:** This large study of prospectively-collected lupus pregnancies demonstrates a decrease in SLE activity among woman who continue HCQ through pregnancy and no harm to pregnancy outcomes. Like all studies of HCQ in lupus pregnancy, this study is confounded by indication and non-adherence. As this study confirms the safety of HCQ and diminished SLE activity with use, it is consistent with current recommendations to continue HCQ throughout pregnancy.

## INTRODUCTION

The guidelines surrounding medication use to treat systemic lupus erythematosus (SLE, lupus) during pregnancy have largely been based on relatively small, University-based, retrospective or prospective cohort studies. Outside of pregnancy, hydroxychloroquine (HCQ) has been demonstrated to decrease lupus flares, risk of lupus nephritis, renal damage from lupus nephritis, and death.[1, 2] HCQ has been prescribed by rheumatologists with expertise in lupus and pregnancy since the early 1990’s, initially based on several small patient series and one small randomized trial showing higher rates of lupus activity and pregnancy complications in the 10 women randomized to stop or not take the drug compared to the 10 who took it throughout pregnancy.[3-6] Analyses of retrospective and prospective lupus pregnancy cohorts also demonstrate some benefits, though generally not as dramatic as in this randomized study. The clearest signal has come from the Hopkins Lupus Cohort, in which women who stopped the drug for pregnancy had significantly higher rates of SLE flare, though not increased rates of pregnancy loss or preterm birth.[7, 8] A recent meta-analysis of published observational studies of HCQ suggested a decrease in preeclampsia and gestational HTN.[9] Early data demonstrated no increase in congenital anomalies in infants with in utero HCQ exposure for malaria and more recent studies have confirmed this finding.[10-12] A recent comparison of pregnancies in the United States with and without a filled HCQ prescription in the first trimester and found a small but statistically significant increase in malformations, though two similar studies in Denmark and Canada did not find an increase in birth defects.[13-15] The prescription of HCQ to manage SLE during pregnancy has been adopted as the standard of care in recent years, with multiple national and international rheumatology guideline groups recommending it for all women with lupus.[16-19]

We hypothesized that prior studies were underpowered to identify improvements in pregnancy outcomes related to HCQ. Therefore, we sought to combine the datasets from multiple prospectively collected lupus pregnancy cohorts to enhance our ability to identify the benefits or risks of HCQ therapy for lupus in pregnancy. Because of the small size of these cohorts, most publications did not present outcomes based on HCQ use, so a traditional meta-analysis based on published odds ratios was not possible. Instead, we completed an Independent Patient Data (IPD) meta-analysis, collecting the datasets from each cohort, re-analyzing each cohort in a similar manner, then combining each cohorts’ results into a meta-analysis.

## METHODS

The meta-analysis was performed in accordance with the PRISMA (Preferred Reporting Items for Systematic Reviews and Meta-Analyses) guidelines.[20] The protocol was registered in September 2015 (25938 PROSPERO). Patients were not directly involved in the design, conduct, or reporting of this research.

### Literature Search Strategy

The original literature search was conducted by a librarian and included Medline (via PubMed®), Embase®, the Cochrane Database of Systematic Reviews and Web of Science’s Core Collection (see Supplementary Table). Search terms covered multiple forms of lupus and terms for pregnancies published after 2000. The search identified 2811 potential manuscripts and conference abstracts (Figure 1).

**Figure 1:**
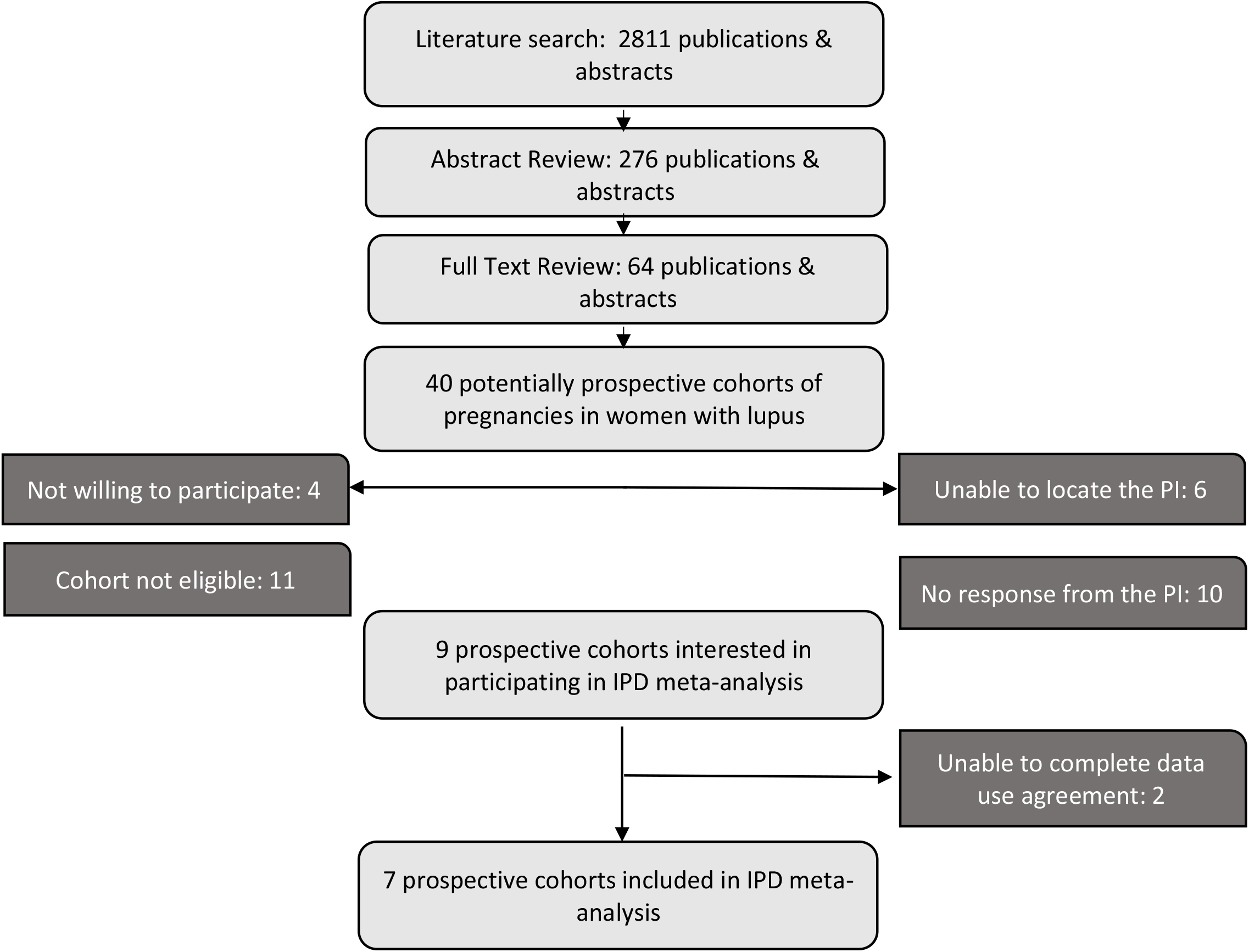
Flow Diagram of Search to identify Eligible Cohorts.

### Cohort Inclusion and Exclusion Criteria

Only prospectively collected cohorts were included to avoid the bias of retrospective assignment of lupus activity (Table 1). All women provided informed consent prior to enrollment in each cohort study, either prior to conception or prior to the completion of the pregnancy. All cohorts had to have data about HCQ use, lupus activity, and pregnancy outcomes. To participate, the cohort investigator had to enter into a data use agreement with Duke University.

**Table 1:** Inclusion Criteria for Cohorts in the Individual Patient Data Meta-analysis:

|  |
| --- |
| **Inclusion Criteria** |
| **Cohort Study Parameters:** |
| Prospective pregnancy data collection |
| Women diagnosed with systemic lupus erythematosus according to a standard set of criteria. |
| Women in the study signed informed consent for participation through an IRB-approved protocol |
| **Features of the Cohort Dataset** |
| Includes an assessment of lupus activity |
| Includes medications used to treat lupus during pregnancy |
| Includes information about pregnancy outcomes |
| **Features of the Publication** |
| Manuscripts published between 2000-2015 |
| Abstracts published between 2012-2015 at the ACR or EULAR Scientific Meetings |
| The cohort includes at least 25 pregnancies in women with lupus |
| Manuscript published in English |
| **Data Access – for the studies included in the individual patient data meta-analysis** |
| A member of the original research team is able to provide the clinical data and is willing to discuss the specific measures and data fields within cohort with the meta-analysis team |
| A member of the original research team is willing and able to sign the Data Use Agreement and provide data to Duke University. |

### Analysis

#### Individual Participant Analysis

The exposure of interest was HCQ use during pregnancy, which included women who continued HCQ throughout or started in the first trimester and continued through pregnancy. Women who started HCQ in the 2^nd^ or 3^rd^ trimesters or who stopped HCQ during pregnancy were considered “unexposed.” Outcomes of interest were analyzed separately and included fetal loss (at any point during pregnancy), preterm birth (<37 weeks gestation), early preterm birth (<34 weeks gestation), preeclampsia, and high disease activity during pregnancy.

Lupus activity during pregnancy was assessed using different methods in the cohorts. For this meta-analysis, high disease activity was defined as physician global assessment (PGA) >1, SLEDAI (SLE Disease Activity Index) >4 at any point during pregnancy, or flare per the treating rheumatologist, depending on the data available. PGA is on a scale from 0-3, with 0 indicating no lupus activity and 3 indicating severe lupus activity. SLEDAI is a weighted score that attributes varying levels of points to different disease manifestations.[21] A variation for pregnancy, the SLEPDAI, was used in some cohorts.[22] To facilitate the analysis, we generated new variables in each dataset to identify the disease activity, medications, and pregnancy outcomes in a semi-uniform fashion.

All prospective pregnancy cohorts are limited in their ability to collect early pregnancy losses that occur prior to study enrollment. The risk of pregnancy loss decreases with each passing week of gestation, so late enrollment artificially decreases the frequency of pregnancy loss. To limit for this bias, we only included women who were enrolled in their first trimester, prior to gestational week 14. Each cohort was analyzed separately, then the odds ratios for each outcome of interest were combined to determine the overall association of HCQ treatment on pregnancy outcomes. Due to some women having multiple pregnancies within the same cohort, we randomly selected one pregnancy per woman to be included in the analysis. This corrected for any correlation between pregnancy outcomes within a patient. As a sensitivity analysis for the outcome of fetal loss, we excluded any losses that occurred prior to 10 weeks. Twin pregnancies were not included in analyses for the outcomes of preterm birth, early preterm birth, and preeclampsia.

We performed meta-analysis summaries using a DerSimonian-Laird random-effects model and conservatively used the Knapp-Hartung approach to adjust the standard errors of the estimated model coefficients.[23, 24] Binary outcomes were pooled as an odds ratio (OR) with 95% CI which indicates no statistically significant effect when overlapping one. The meta-analyses were performed in R (version 3.5.3) using the “metafor” package (version 2.0-0).[25, 26]

### Heterogeneity of Treatment Effect

Due to the effects of lupus disease severity and manifestations on pregnancy outcomes, the results were estimated for the overall effect, as well as stratified by history of lupus nephritis, APS, and disease activity in the 1^st^ trimester as recorded within each dataset. As we did not have data available on pre-pregnancy disease activity, we addressed bias by indication by presenting results for patients with lupus nephritis, based on the assumption that HCQ prescription would be the standard of care for a patient with a history of or active LN during pregnancy.

### Confounders

Based on a Directed Acyclic Graph (Figure 2), the minimal sufficient adjustment set for the association of HCQ use and pregnancy outcomes includes history of lupus nephritis, disease activity, and year of pregnancy. To account for these confounders, we present results stratified by history of LN, as well as disease activity in the 1^st^ trimester. As we are restricting our analysis to pregnancies since 1995, we have not included year in our model. We did not adjust for prednisone use because it is a causal intermediate between HCQ and pregnancy outcomes and not a confounder; adjusting for prednisone would bias the effect estimate.

**Figure 2:**
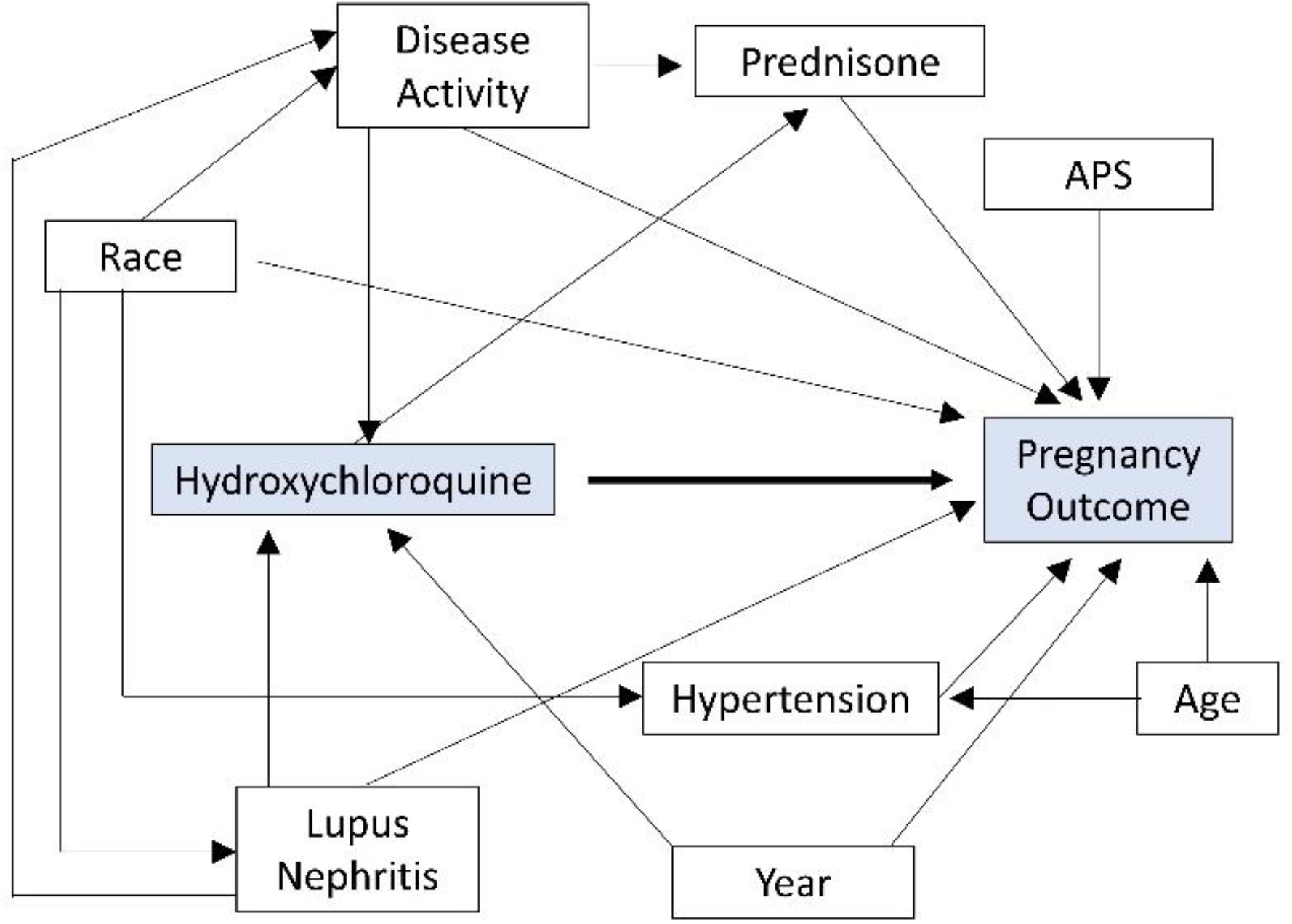
DAG: Directed Acyclic Graph demonstrating the influence of confounders on HCQ use and pregnancy outcomes. Adjustments were made for disease activity and lupus nephritis, as both influence HCQ prescription and pregnancy outcomes. While hypertension, race, and maternal age impact pregnancy outcomes, they do not strongly influence HCQ prescription, so adjustment is not required in this analysis. Year of pregnancy could impact both HCQ prescription and pregnancy outcomes based on changes in practice over time, however the lead physicians in each study did not significantly alter HCQ prescribing habits during the study period so this was excluded from the analysis.

### Risk of Bias Assessment

The study was assessed for bias by the study team using the Risk Of Bias In Non-randomized Studies of Interventions (ROBINS-I).[27]

## RESULTS

### Study selection

Once the inclusion and exclusion criteria were applied, we identified 40 potential cohorts which ultimately resulted in 9 investigators eligible and interested in participating and 7 who were able to contribute datasets (Figure 1). The most common reasons for lack of participation was study ineligibility due to either lack of consent or retrospective data collection (n=11), followed by investigators who did not respond to repeated inquiry (n=10).

### Study characteristics

The 7 cohorts were all collected prospectively through specialized university rheumatology clinics. A total of 962 pregnancies in 821 women were obtained from 7 cohorts; 761 (79%) of pregnancies in 668 women had a 1^st^ trimester visit and the frequency of 1^st^ trimester visits ranged between 61% at Duke University to 100% in Israel (Table 2).

**Table 2:**
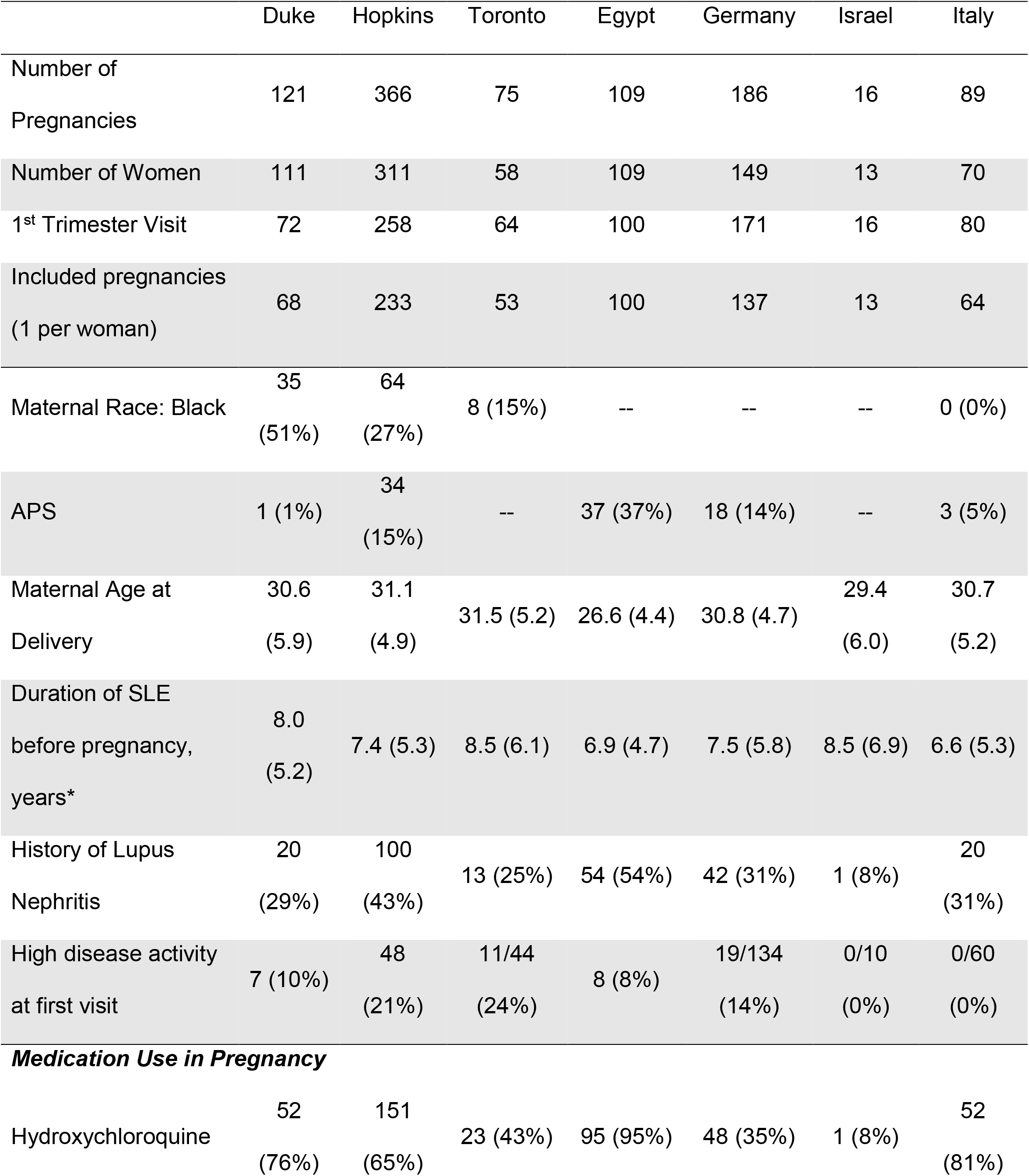
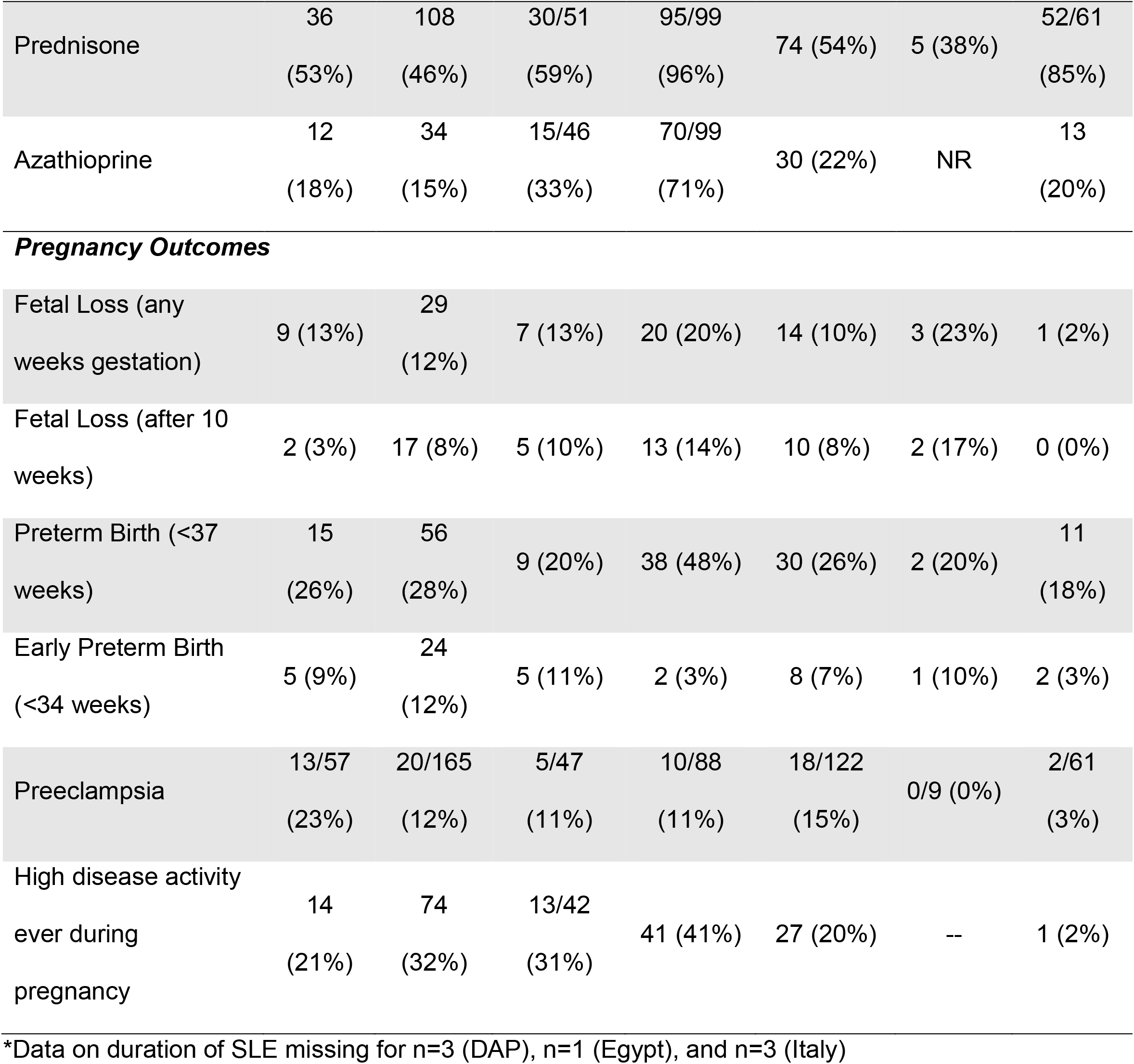
Cohort Characteristics

|  | Duke | Hopkins | Toronto | Egypt | Germany | Israel | Italy |
| --- | --- | --- | --- | --- | --- | --- | --- |
| Number of Pregnancies | 121 | 366 | 75 | 109 | 186 | 16 | 89 |
| Number of Women | 111 | 311 | 58 | 109 | 149 | 13 | 70 |
| 1<sup>st</sup> Trimester Visit | 72 | 258 | 64 | 100 | 171 | 16 | 80 |
| Included pregnancies<br>(1 per woman) | 68 | 233 | 53 | 100 | 137 | 13 | 64 |
| Maternal Race: Black | 35<br>(51%) | 64<br>(27%) | 8 (15%) | -- | -- | -- | 0 (0%) |
| APS | 1 (1%) | 34<br>(15%) | -- | 37 (37%) | 18 (14%) | -- | 3 (5%) |
| Maternal Age at<br>Delivery | 30.6<br>(5.9) | 31.1<br>(4.9) | 31.5 (5.2) | 26.6 (4.4) | 30.8 (4.7) | 29.4<br>(6.0) | 30.7<br>(5.2) |
| Duration of SLE<br>before pregnancy,<br>years* | 8.0<br>(5.2) | 7.4 (5.3) | 8.5 (6.1) | 6.9 (4.7) | 7.5 (5.8) | 8.5 (6.9) | 6.6 (5.3) |
| History of Lupus<br>Nephritis | 20<br>(29%) | 100<br>(43%) | 13 (25%) | 54 (54%) | 42 (31%) | 1 (8%) | 20<br>(31%) |
| High disease activity<br>at first visit | 7 (10%) | 48<br>(21%) | 11/44<br>(24%) | 8 (8%) | 19/134<br>(14%) | 0/10<br>(0%) | 0/60<br>(0%) |
| **Medication Use in Pregnancy** |  |  |  |  |  |  |  |
| Hydroxychloroquine | 52<br>(76%) | 151<br>(65%) | 23 (43%) | 95 (95%) | 48 (35%) | 1 (8%) | 52<br>(81%) |
| Prednisone | 36<br>(53%) | 108<br>(46%) | 30/51<br>(59%) | 95/99<br>(96%) | 74 (54%) | 5 (38%) | 52/61<br>(85%) |
| Azathioprine | 12<br>(18%) | 34<br>(15%) | 15/46<br>(33%) | 70/99<br>(71%) | 30 (22%) | NR | 13<br>(20%) |
| ***Pregnancy Outcomes*** |  |  |  |  |  |  |  |
| Fetal Loss (any weeks gestation) | 9 (13%) | 29<br>(12%) | 7 (13%) | 20 (20%) | 14 (10%) | 3 (23%) | 1 (2%) |
| Fetal Loss (after 10 weeks) | 2 (3%) | 17 (8%) | 5 (10%) | 13 (14%) | 10 (8%) | 2 (17%) | 0 (0%) |
| Preterm Birth (<37 weeks) | 15<br>(26%) | 56<br>(28%) | 9 (20%) | 38 (48%) | 30 (26%) | 2 (20%) | 11<br>(18%) |
| Early Preterm Birth (<34 weeks) | 5 (9%) | 24<br>(12%) | 5 (11%) | 2 (3%) | 8 (7%) | 1 (10%) | 2 (3%) |
| Preeclampsia | 13/57<br>(23%) | 20/165<br>(12%) | 5/47<br>(11%) | 10/88<br>(11%) | 18/122<br>(15%) | 0/9 (0%) | 2/61<br>(3%) |
| High disease activity ever during pregnancy | 14<br>(21%) | 74<br>(32%) | 13/42<br>(31%) | 41 (41%) | 27 (20%) | -- | 1 (2%) |

\*Data on duration of SLE missing for n=3 (DAP), n=1 (Egypt), and n=3 (Italy)

There were important variations between the women’s characteristics, medications prescribed, and pregnancy outcomes between the cohorts. Race was recorded in 4 cohorts, the frequency of antiphospholipid syndrome (APS) varied widely and a one-third of all pregnancies occurred in a woman with a history of lupus nephritis.

HCQ was the most commonly prescribed medication, with the majority of women taking this drug throughout pregnancy. The Israel, Toronto, and Germany cohorts had fewer than 50% of women on this medication. Prednisone was the next most common prescription followed by azathioprine.

Pregnancy loss occurred in 12%, ranging from 2% in Italy to 23% in Israel. Preterm birth occurred in 28% of all live births and preeclampsia in an estimated 12%. High SLE activity was present in 26% of all pregnancies, but just 2% of Italian pregnancies and 41% of Egyptian pregnancies.

### IPD integrity

Missing data, in particular missing pregnancy outcomes, medications, and disease activity, significantly decreased the number of pregnancies in the cohorts that were available for study.

### Bias Assessment

Using the Risk of Bias in Non-randomized studies of Inventions (ROBINS-I) method, the overall study has a serious risk of bias, driven by confounding by indication and the unknown, but likely significant, frequency for non-adherence with HCQ.

## Results

HCQ decreased the risk of high disease activity during pregnancy among women taking HCQ (OR: 0.53; 95% CI: 0.31, 0.93). HCQ did not, however, impact the pregnancy outcomes of fetal loss, preterm delivery or preeclampsia in the overall population of pregnancies in women with lupus (Table 3; Figure 3).

**Table 3.** Summary of pooled odds ratios for the association of HCQ with pregnancy outcomes among women with lupus. Sensitivity analyses stratified pregnancies by 1^st^ trimester SLE activity, whether the woman had a history of lupus nephritis, and whether the woman had a history of antiphospholipid syndrome (APS)

|  | N | Fetal Loss<br>after 10 wks | Preterm Birth | Preeclampsia | High SLE<br>Activity |
| --- | --- | --- | --- | --- | --- |
| All SLE pregnancies | 668 | 0.83<br>(0.63, 1.09) | 0.68<br>(0.39, 1.18) | 0.89<br>(0.62, 1.29) | 0.53<br>(0.31, 0.93) |
| Low SLE Activity<br>1<sup>st</sup> trim. | 557 | 1.00<br>(0.62, 1.60) | 0.55<br>(0.30, 0.99) | 0.77<br>(0.43, 1.40) | 0.56<br>(0.25, 1.22) |
| High SLE Activity<br>1<sup>st</sup> trim. | 93 | 0.73<br>(0.09, 6.15) | 1.03<br>(0.43, 2.46) | 1.73<br>(0.23, 13.00) | -- |
| Lupus nephritis<br>History | 250 | 0.59<br>(0.14, 2.50) | 0.67<br>(0.33, 1.37) | 0.65<br>(0.16, 2.63) | 0.49<br>(0.27, 0.91) |
| No lupus<br>nephritis History | 418 | 1.08<br>(0.46, 2.57) | 0.75<br>(0.34, 1.68) | 0.94<br>(0.48, 1.83) | 0.61<br>(0.32, 1.18) |
| APS | 93 | 0.46<br>(0.10, 2.06) | 0.88<br>(0.09, 8.97) | 0.60<br>(0.28, 1.28) | 0.91<br>(0.06, 13.63) |
| No APS | 509 | 1.01<br>(0.42, 2.41) | 0.80<br>(0.25, 2.58) | 1.03<br>(0.70, 1.51) | 0.58<br>(0.41, 0.81) |

**Figure 3.**
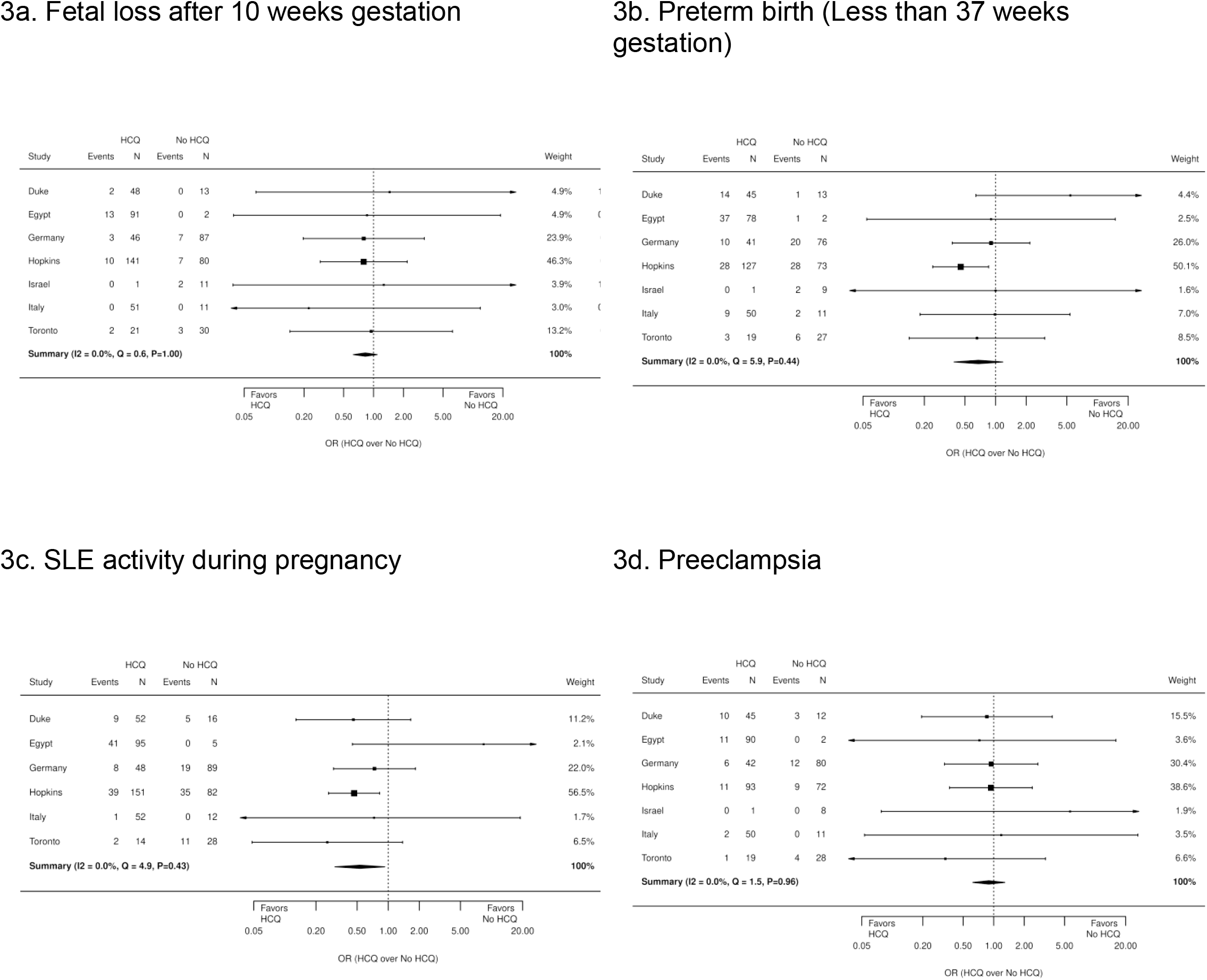
Forest plots for the effect of HCQ use on pregnancy outcomes in women with SLE.

### Subgroup Analysis

When the analysis was stratified by disease activity in the 1^st^ trimesters, those with low disease activity early in pregnancy who took HCQ had a decreased risk of preterm delivery (OR: 0.55; 95% CI: 0.30, 0.99). On the other hand, HCQ use did not appear to impact the rate of preterm birth among women with more active SLE in the first trimester. Whether a woman had high lupus activity in her first trimester or not, HCQ did not impact the risk for fetal loss or preeclampsia.

Results were similar when the analysis was restricted to women with prior or current lupus nephritis. High disease activity was reduced among women with a history of lupus nephritis taking HCQ, but HCQ was not significantly related with fetal loss, preterm birth, or preeclampsia in women with or without a nephritis.

Among women with and without APS HCQ remained unrelated to fetal loss, preterm birth, or preeclampsia, but there was a decreased risk of high disease activity during pregnancy among women without APS.

A reanalysis using a different random selection of pregnancies (one per woman) produced the same results, with no significant association between HCQ use and fetal loss, preterm delivery, or preeclampsia but a decreased risk for high disease activity in women taking HCQ.

## DISCUSSION

By combining datasets from seven prospective lupus pregnancy cohorts in an independent patient data meta-analysis we found that taking HCQ throughout pregnancy decreased the risk for high SLE activity in pregnancy and had no impact on fetal loss, preterm delivery, or preeclampsia. Among women with low SLE activity early in pregnancy, taking HCQ was associated with a lower risk for preterm birth; this benefit was not found in women with highly active SLE. Three prior systematic reviews and/or meta-analyses including a range of studies, though none using a patient-by-patient analysis as we have employed, each reached a similar conclusion: HCQ did not significantly impact pregnancy outcomes.[28-30] Specifically, these studies noted that prematurity, intrauterine growth restriction (IUGR), congenital malformations, low birth weight, and stillbirth were not associated with HCQ. Only one analysis identified a higher risk of early pregnancy loss in women with HCQ, but this was not confirmed in another analysis, nor in this analysis. While we were not able to include data from the PROMISSE study, the results of this multi-center prospective study were similar, with no difference in the frequency of adverse pregnancy outcomes in women with and without HCQ use.[31] Several recent studies have suggested pregnancy benefits from HCQ beyond SLE. Three recent studies suggested that HCQ may play a role in improving pregnancy outcomes for women with APS; we did not see a benefit from HCQ in 97 women with both SLE and APS.[32-37] A clinical trial is currently underway testing the efficacy of HCQ in women with refractory APS.[38] One retrospective study and a meta-analysis demonstrated a decrease in preeclampsia among women with lupus taking HCQ in pregnancy; we did not find a similar benefit studying 65 SLE pregnancies with preeclampsia, the largest dataset available.[9, 39] Finally, HCQ has been demonstrated in several retrospective studies and one prospective study to significantly decrease the incidence of congenital heart block (CHB) due to maternal Ro/SSA antibodies; this study did not include a sufficient number of CHB cases to study this association.[40-44]

This meta-analysis does not address congenital defects or long-term infant outcomes because the cohorts were not designed for systematic ascertainment of congenital defects nor the infant outcomes beyond the several weeks after delivery. Several recent studies, however, have assessed the risk of congenital defects with HCQ or chloroquine exposure in large administrative databases with varying results. While a large study of two US national databases found a significant increase in birth defects, no increase was found in similar studies from Denmark, Quebec, two US state-based datasets, or an Israeli teratology study.[12-15, 45] Ocular toxicity from *in utero* exposure has not been identified in 12 cohorts and randomized trials.[46]

An important strength of this paper is that, by having access to the individual patient-level data, we were able to run similar analyses on each dataset so that our analysis was not restricted to previously published study results. A key strength of this project is the prospective collection of pregnancies, with disease activity and medications recorded prior to delivery. Additionally, all pregnancies were enrolled in the first trimester and managed by a rheumatologist with a particular interest in pregnancy management, suggesting that, within each center, patients received similar, state-of-the art care with or without HCQ therapy. By including multiple centers, this study incorporates pregnancies from across the globe and enhances the generalizability of the study to multiple races and ethnicities of patient.

Despite reporting on the largest, detailed, prospectively-collected set of lupus pregnancies, this study contains multiple potential biases that confound the results and conclusions. As pregnancies occurred over a 20-year period and in 7 different centers, all pregnancies did not receive uniform obstetric or rheumatologic care. While the majority of women were prescribed 400mg of HCQ each day, the exact dose throughout pregnancy is not known. As HCQ is often weight-based, some women may have been under-dosed, particularly in light of weight gain during pregnancy.[47] Bias by indication may have obscured clinical benefits of HCQ. All care was ‘standard of care’ and determined at the discretion of the treating physicians and pregnant women; women were not randomized to HCQ. Women with mild lupus and the lowest risk for pregnancy complications may have been less likely to be prescribed HCQ, thus improving the pregnancy outcomes for the non-HCQ group. In a sub-analysis women with either current or prior lupus nephritis, who likely all have a uniform indication for HCQ; HCQ was not associated with significantly different pregnancy outcomes. Missing data points were not unusual as each of these studies was collected as an adjunct to clinical care and led to the need to exclude some pregnancies from the analysis. We hypothesize that missing data was not entirely random with higher-risk pregnancies more likely to have delayed presentation for care, more missed appointments, and more incomplete data. Each cohort was designed independently and collected different data, preventing adjustment for a broad range of confounders.

Our study could not account for adherence and it is likely that some women who did not actually take the drug were included in the HCQ group. Outside of pregnancy, 18-44% of patients with SLE who report taking HCQ are not taking it based on blood HCQ levels.[48-50] In 25 women with SLE within the DAP Registry (Duke University), 24% of women who reported taking HCQ had levels demonstrating non-adherence; preterm delivery occurred in 83% of non-adherent mothers compared to 21% of adherent mothers (p<0.01).[51] Based on these prior studies, we estimate that about a quarter of women in the HCQ group of each cohort may be non-adherent and are thus miss-classified in the HCQ group. Given the size of this variability, this miss-classification could obscure any clinical benefit from HCQ. Overcoming this bias at this point is not possible, but future prospective studies should include measurements of HCQ levels to better classify medication use.

In summary, this is the largest, prospective study of SLE pregnancy, including 7 cohorts with over 900 pregnancies collected world-wide over the last several decades. Using an independent patient-level meta-analysis we found that HCQ use throughout pregnancy was associated with decreased SLE activity, but was not associated with pregnancy loss, preterm birth, preeclampsia. Among women with low SLE activity in the first trimester, however, HCQ was associated with lower rates of preterm birth. Given the near ubiquity of HCQ therapy in young women with SLE and the demonstrated benefit in lupus activity from continued HCQ in pregnancy, the finding that continuing HCQ in pregnancy does not negatively impact pregnancies is reassuring. Potential biases of indication, missing data, and non-adherence are serious limitations to this and all currently available data on HCQ use in lupus pregnancy. These limitations can only be overcome through a large, randomized, double-blind trail of HCQ in lupus pregnancy that includes frequent measurements of HCQ levels. Taken together, this study supports the current recommendations from the American College of Rheumatology, EULAR, British Rheumatology Association to continue HCQ during lupus pregnancy.[16, 18, 19, 52]

## Data Availability

All data produced in the present study are available upon reasonable request to the authors.

## Acknowledgements and Affiliations

NA

## Funding

Clowse: AHRQ 1K18HS023443-01A1; Arthritis Foundation, Arthritis Investigator Award. Industry affiliation: Grant and consultant fees from GSK.

Eudy: NIH NCATS Award Number 1KL2TR002554

Petri: NIH RO-1 AR069572

## Key Messages

- Multiple national and international guidelines for the management of SLE in pregnancy currently recommend the use of hydroxychloroquine throughout pregnancy.
- This is the largest study to date of prospectively-collected data from women with SLE throughout pregnancy, combining over 900 pregnancies from 7 studies from across the world.
- This independent patient data meta-analysis found that the prescription of HCQ is associated with decreased SLE activity in pregnancy, however it does not demonstrate a significant improvement in pregnancy outcomes.
- This study demonstrates that observational data, which is the best existing data on pregnancy management for women with rheumatic diseases, is insufficient to clarify the risks and benefits of HCQ.
- Controlled trials without the biases of indication, missing data, and unmeasured non-adherence are essential to understand the impact of HCQ on SLE pregnancy outcomes.

## Supplementary Table 1: Literature Search

### Literature search strategy

The original literature search was conducted by a master’s level librarian in the Duke University Medical Library. It included a search of PubMed^®^, Embase^®^, and the Cochrane Database of Systematic Reviews. For PubMed, lupus manuscripts were identified using the Mesh terms: lupus erythematosus, systemic; lupus coagulation inhibitor; lupus vasculitis, central nervous system; lupus erythematosus, discoid; lupus erythematosus, cutaneous; neonatal systemic lupus erythematosus’ lupus vulgaris; and lupus as a title or abstract word. Pregnancy manuscripts were identified using the Mesh terms pregnancy, pregnant women, and prenatal care; the title/abstract words were pregnancy, pregnant, pregnancies, and gestation. These searches were combined and animal studies, non-English studies, studies prior to 2000, and case reports were removed. This search identified 1342 manuscripts (Supplementary Table 1).

The Cochrane search used lupus in the title, abstract or keywords and identified pregnancy using pregnancy, pregnant women, pregnant, pregnancies, gestation, and prenatal care. When combined, this search identified 56 manuscripts. The EMBASE search identified lupus manuscripts with the following expressions lupus vulgaris, lupus anticoagulant, lupus erythematosus, lupus erythematosus, systemic, lupus coagulation inhibitor, lupus vasculitis, central nervous system; lupus erythematosus, discoid; lupus erythematosus, cutaneous; neonatal systemic lupus erythematosus. Pregnancies were identified using the expressions, titles, and abstract terms of pregnancy, pregnant woman, prenatal care, pregnant, pregnancies, gestation, and prenatal care. When combined and non-human, non-English, manuscripts prior to 2000, and case reports were excluded, 1712 manuscripts were identified. Another 552 conference abstracts were then added, resulting in 2264 possible studies. Web of Science was also queried. The topic lupus was combined with pregnancy, using the same list of terms as above. This resulted in 2025 possible studies. Once the duplicate studies were removed from these searches, we had a total of 2811 manuscripts and conference abstracts to review.

| Search | Query | Items found |
| --- | --- | --- |
| #1 | Search ("Lupus Erythematosus, Systemic"[Mesh] OR "Lupus Coagulation Inhibitor"[Mesh] OR "Lupus Vasculitis, Central Nervous System"[Mesh] OR "Lupus Erythematosus, Discoid"[Mesh] OR "Lupus Erythematosus, Cutaneous"[Mesh] OR "Neonatal Systemic lupus erythematosus" [Supplementary Concept] OR "lupus vulgaris"[MeSH Terms] OR "lupus"[tiab]) | 73398 |
| #2 | Search "pregnancy"[MeSH Terms] OR "Pregnant Women"[Mesh] OR "pregnancy"[tiab] OR "pregnant"[tiab] OR "pregnancies"[tiab] OR "gestation"[tiab] OR "Prenatal Care"[Mesh] | 825724 |
| #3 | Search (#1 AND #2) | 4039 |
| #4 | Search #3 NOT ("Animals"[MeSH] NOT "Humans"[Mesh]) | 3968 |
| #5 | Search #4 Filters: English | 3151 |
| #6 | Search #5 Filters: Publication date from 2000/01/01; English | 1615 |
| #7 | Search #6 NOT ("Case Reports" [Publication Type] OR "a case report"[ti] OR ": case report"[ti]) | 1342 |

Cochrane:
| ID | Search | Hits |
| --- | --- | --- |
| #1 | pregnancy OR Pregnant Women OR pregnancy OR pregnant OR pregnancies OR gestation OR Prenatal Care | 29482 |
| #2 | Lupus:ti,ab,kw (Word variations have been searched) | 1171 |
| #3 | #1 and #2 | 56 |

EMBASE:
| No. | Query | Results |
| --- | --- | --- |
| #1 | 'lupus vulgaris'/exp OR 'lupus anticoagulant'/exp OR 'lupus erythematosus'/exp OR 'lupus erythematosus, systemic' OR 'lupus coagulation inhibitor' OR 'lupus vasculitis, central nervous system' OR 'lupus erythematosus, discoid' OR 'lupus erythematosus, cutaneous' OR 'neonatal systemic lupus erythematosus':ab,ti OR 'lupus vulgaris' OR 'lupus':ab,ti | 111333 |
| #2 | 'pregnancy'/exp OR 'pregnant woman'/exp OR 'prenatal care'/exp OR<br>'pregnancy':ti,ab OR 'pregnant':ti,ab OR 'pregnancies':ti,ab OR<br>'gestation':ti,ab OR 'Prenatal Care':ti,ab | 825724 |
| #3 | #1 AND #2 | 5886 |
| #4 | #3 AND [humans]/lim AND [english]/lim AND [2000-2015]/py | 3005 |
| #5 | #4 AND (([article]/lim OR [article in press]/lim OR [review]/lim) NOT ('case<br>report'/exp OR 'a case report':ti OR ': case report':ti)) | 1712 |
| #6 | #3 AND ([conference abstract]/lim OR [conference paper]/lim) AND<br>[humans]/lim AND [english]/lim AND [2012-2015]/py | 552 |
| #7 | #5 OR #6 | 2264 |

Web of Science:
| Set | History | Results |
| --- | --- | --- |
| #1 | TOPIC: ((pregnancy OR Pregnant Women OR pregnancy OR pregnant<br>OR pregnancies OR gestation OR "Prenatal Care")) AND TOPIC: (lupus)<br>Refined by: DOCUMENT TYPES: (PROCEEDINGS PAPER OR<br>MEETING ABSTRACT) AND PUBLICATION YEARS: (2012-2015) | 65 |
| #2 | (TI=(lupus AND pregnancy)) AND LANGUAGE: (English)<br>Refined by: DOCUMENT TYPES: (ARTICLE OR REVIEW) AND<br>PUBLICATION YEARS: (2000-2015) | 331 |
| #3 | #1 OR #2 | 2,025 |

## Notes

Potential Conflicts of Interest: This manuscript was not funded by any commercial entities.

No other authors reports conflicts of interest

### Competing Interest Statement

Clowse: Funding: AHRQ 1K18HS023443-01A1, Arthritis Foundation; Grants: GSK; Consulting and Advisory Board: UCB. Eudy: Grant funding: NIH NCATS Award 1KL2TR002554. Balevic: Grants: NIH, US FDA, PCORI, RRF, CARRA; Consulting: UCB; Travel: FDA; Leadership: CARRA Assistant Scientific Director. Gladman: Grants and or consulting fees from AstraZeneca; Leadership: Member of the Systemic Lupus International Collaborating Clinics group; served on the Medical advisory board of the Lupus foundation of America. Petri: Grant NIH RO-1 AR069572. The other authors report no significant competing interests or grant funding that pertains to the research within this manuscript.

### Funding Statement

Clowse: AHRQ 1K18HS023443-01A1; Arthritis Foundation, Arthritis Investigator Award. Eudy: NIH NCATS Award Number 1KL2TR002554 Petri: NIH RO-1 AR069572

### Author Declarations

Each dataset was collected under a local IRB protocol and patients provided informed consent prior to participation. The meta-analysis was approved by Duke University IRB; protocol number Pro00066961.

